# Alternative Factor Prescribing after Low-Dose Recombinant Factor VIIa Protocol in Cardiac Surgery

**DOI:** 10.1101/2022.04.30.22274528

**Authors:** Amanda Momenzadeh, Jesse G. Meyer, Noelle De Leon, Candy Tsourounis

## Abstract

**Background:** Safety concerns exist with the off-label use of recombinant factor VIIa (rFVIIa, Novoseven RT®) for refractory bleeding in cardiac surgery, including increased risk of thromboembolism. A rFVIIa protocol was implemented in December 2015 to standardize rFVIIa for cardiac surgery related hemorrhage.

**Methods:** We performed a retrospective, observational review of rFVIIa in adult cardiac surgery patients pre-protocol (January 2015 to November 2015) vs. post-protocol (December 2015 to March 2016). Study outcomes were rate of rFVIIa administration, rFVIIa dosing characteristics, length of stay, mortality, readmission rate, need for re-exploration, and rate of 4-factor Prothrombin Complex Concentrates (PCC; Kcentra®) administration.

**Results:** There was a significant reduction in percentage of cardiac surgery cases receiving rFVIIa pre-vs. post-protocol (14.3 vs. 5.2%, p=0.015). Average total dose per patient decreased between groups (81.4 vs. 56.6 mcg/kg, p=0.059). In-hospital mortality, length of stay, need for re-exploration, readmission rates and 30-day mortality did not differ. Although 4-four-factor PCC significantly increased post-protocol (2.5% vs. 8%, p=0.02), overall use of factor products, rFVIIa or 4-factor PCC, did not change between study periods (16.8% vs. 13%, p=0.416). Mean cost of either rFVIIa or 4-factor PCC pre-protocol was significantly higher than that post-protocol ($8,778 vs. $4,421, p=0.0008).

**Conclusions:** The use of rFVIIa decreased after implementation of a rFVIIa protocol targeting 30 mcg/kg/dose without compromising morbidity or mortality outcomes. Four-factor PCC use significantly increased during the study, but the overall cost was reduced. Institutions wanting to implement a rFVIIa protocol should take careful measures to concurrently address off-label use of 4-factor PCC.

## Introduction

The management of life-threatening bleeding related to cardiac surgery remains a critical problem. Post-operative bleeding is associated with increased morbidity and mortality,^1^ and has been documented to occur in 5–15% of cardiac surgical patients with cardiopulmonary bypass (CPB).^2^ Recombinant factor VIIa (rFVIIa, Novoseven RT®) is a hemostatic agent that binds to the surface of activated platelets and tissue factor released from the subendothelium at the site of injury.^3^ This process results in factor X activation, converting prothrombin into thrombin, which ultimately leads to the formation of a fibrin meshwork.^3^ Although rFVIIa’s half-life is less than six hours, this burst of thrombin continues to be active past the peri-operative period due to the half-life of factor II, which is up to 72 hours.^4^ rFVIIa is FDA-approved for the management of bleeding in adults and children with hemophilia A and B with inhibitors, congenital factor VII deficiency, Glanzmann’s thrombasthenia, and acquired hemophilia.^5^ In practice, rFVIIa is commonly used off-label for hemorrhage during cardiac surgery. Between 2000 and 2008, the inpatient rFVIIa off-label use increased by more than 140-fold nationwide, compared to a 4-fold increase in patients with hemophilia.^6^ In 2008, 97% of rFVIIa use was for off-label purposes, 29% of which consisted of adult and pediatric cardiovascular surgery.^6^

To date, no study has demonstrated a definitive mortality benefit with rFVIIa for any off-label indication. This is largely because the available studies are underpowered and difficult to interpret due to significant heterogeneity in the patients studied and the endpoints measured. In 2005, the prescribing information for rFVIIa was modified to include a warning of serious arterial and venous thrombotic events associated with off-label use. ^3^ A 2010 systematic review evaluated the rate of TE events within 35 randomized clinical trials, 6% of whom included cardiac surgery patients that received rFVIIa, and found a significantly higher rate of arterial TE events in patients who received rFVIIa than in those who received placebo (5.5% vs. 3.2%, p = 0.003).^7^ A 2012 Cochrane review of 29 randomized controlled trials including prophylactic and therapeutic rFVIIa use found no evidence of mortality benefit and a trend indicating an increased risk of thromboembolic (TE) events with rFVIIa.^8^ In 2014, a retrospective, five-year study including 28,000 cardiac surgery patients with perioperative hemorrhage reported an in-hospital mortality frequency of 40% among those treated with rFVIIa compared to 18% in control patients (p<0.001).^9^ Taken together, a clear safety concern persists for off-target thrombosis risk associated with rFVIIa in clotting independent of tissue factor.^3^

Despite the concerning safety data published on the use of rFVIIa in cardiothoracic (CT) surgery patients without hemophilia, the 2011 and 2019 Society of Cardiovascular Anesthesiologists Clinical Practice Improvement Advisory for Management of Perioperative Bleeding and Hemostasis in Cardiac Surgery Patients recommend use of rFVIIa in open heart surgical patients with bleeding refractory to routine hemostatic agents (Class IIb recommendation; level of evidence B).^10,11^ The guideline is partly based on a pivotal 2009 randomized, placebo-controlled trial (30 sites) evaluating the efficacy and safety of rFVIIa in patients with bleeding greater than 200 mL/hour post cardiac surgery.^12^ Critically serious adverse events were greater but did not reach statistical significance in the rFVIIa group. ^6^ Rates of reoperation, blood loss, and transfusion were all significantly reduced in the rFVIIa group compared to placebo.^12^ Importantly, this study is commonly used to justify the off-label use of rFVIIa for the management of bleeding during cardiac surgery.^13^

Due to the heterogeneity in rFVIIa dosing used across studies, a safe and effective dosing schedule has been difficult to determine. The approved dose in hemophilia A and B with inhibitors is 90 mcg/kg and doses in studies using rFVIIa for surgical bleeding in cardiac surgery have ranged widely. The literature indicates no difference in efficacy endpoints between low doses (<40 mcg/kg) compared to medium doses (41-80 mcg) and high doses (81-100 mcg/kg) of rFVIIa.^9,14–22^ Even lower doses of rFVIIa (range 11.1-32 mcg/kg) have been reported as efficacious. ^9,14–21,23,24^ Low doses of rFVIIa have not been associated with increased adverse events, while higher doses of rFVIIa have been shown to significantly increase the risk of arterial TE events.^9,14–21,23,24^ Given lower doses have demonstrated similar efficacy in controlling bleeding and a favorable safety profile, lower doses of rFVIIa should be preferred during CT surgery. Given the safety concerns with the off-label use of rFVIIa, a pharmacist-driven, interdisciplinary initiative was developed to standardize dosing for microvascular bleeding in cardiac surgery. Through the efforts of an interdisciplinary team of cardiac surgeons, anesthesiologists, nurses, and pharmacists, a rFVIIa dosing protocol was implemented at the University of California, San Francisco (UCSF) Medical Center in December 2015. Few studies have documented the effects of institutional protocols aimed at restricting the use and dose of rFVIIa for cardiac surgery. The objective of this study was to determine the effect of a rFVIIa stewardship protocol targeting 30 mcg/kg dosing on prescribing practices, patient outcomes, and cost at a large academic medical center.

## Methods

### Institutional Guideline

UCSF Medical Center is a 690-bed tertiary academic medical center with a robust adult cardiothoracic surgery division, which specializes in cardiac surgery, general thoracic surgery and heart and lung transplant. An interdisciplinary group at UCSF Medical Center consisting of cardiac surgeons, cardiac anesthesiologists, nurses and pharmacists developed a rFVIIa weight-based rFVIIa dosing protocol in cardiac surgery patients, which was approved by the Hemostasis and Antithrombic Committee in October 2015, and the Pharmacy and Therapeutics Committee in December 2015. The protocol was implemented in December 2015, and education was provided to cardiac intensive care unit (ICU) nurses and critical care and operating room (OR) pharmacists between January and March 2016.

Cardiac surgery patients eligible to receive rFVIIa must have had brisk bleeding or have been categorized as high risk for bleeding (history of coagulopathy or recent anticoagulants, aortic root or dissection, endocarditis, long pump run or clamp time, LVAD/heart transplant, re-do valve or CABG, or any bleed after protamine has been given). Additionally, these patients must have failed an institutional algorithm for management of massive refractory blood loss in the OR and ICU, which includes rotational thromboelastometry (ROTEM) guided administration of platelets, cryoprecipitate, or Fresh Frozen Plasma (FFP). Once these criteria were met, the provider was allowed to order rFVIIa according to weight-based dosing (using actual body weight) outlined in the protocol, which targets 30 mcg/kg/dose. The recommended intravenous bolus dose of rFVIIa was 2 mg for patients weighing less than 100 kg, and 3 mg for patients weighing 100 kg or more. If bleeding was uncontrolled after one dose of rFVIIa, a repeat dose of the same strength was allowed per the surgeon’s discretion.

### Study Design

Data were collected through a retrospective review of electronic medical records of patients hospitalized at UCSF who underwent cardiac surgery between January 1, 2015, and November 30, 2015 (pre-protocol group) and between December 1, 2015, and March 31, 2016 (post-protocol group). Patients included in the study were ≥18 years of age, had undergone cardiac surgery, and received at least one dose of rFVIIa in either the adult operating room or post-operative cardiac ICU setting. Patients with the following characteristics were excluded: hemophilia A or B, von Willebrand Disease, factor II, V, VII, X, or XII deficiency, end-stage liver disease, lupus anticoagulant, or Factor V Leiden. Institutional review board (IRB) approval was obtained for this study through the University of California, San Francisco, Committee on Human Research Protection Program.

### Statistics

Counts and percentages were reported for categorical data and means and standard deviations for continuous variables. Chi-square test with a two-sided alternative hypothesis was used to compare proportions of categorical values. Two-sided Wilcoxon rank-sum test was used for the comparison of continuous, non-normally distributed variables. Statistical significance was based on a two-tailed p-value of ≤ 0.05. All data analysis was performed in Python version 3.7.11.

## Results

### Study Population

Figure 1. outlines the study subject selection process. During the pre-protocol period (January 1, 2015, to November 30, 2015), there were 357 adult cardiac surgery cases that met inclusion criteria, of which 51 received rFVIIa. During the post-protocol period (December 1, 2015 to March 31, 2016), 115 adult cardiac surgery cases were performed that met inclusion criteria, of which 6 received rFVIIa. Baseline characteristics of patients who received rFVIIa between the pre- and post-protocol groups were comparable (**Table 1**). The majority of patients in both groups were male, Caucasian, and underwent a cardiac valve replacement or repair procedure. The average age (57.8 vs. 60.1 years), BMI (25.4 vs. 26.2 kg/m^2^), weight (75.1 vs. 83 kg), and whether patients received anti-fibrinolytic therapy in the operating room, i.e., tranexamic acid (TXA) or aminocaproic acid (AA), were not different between groups.

**Figure 1.**
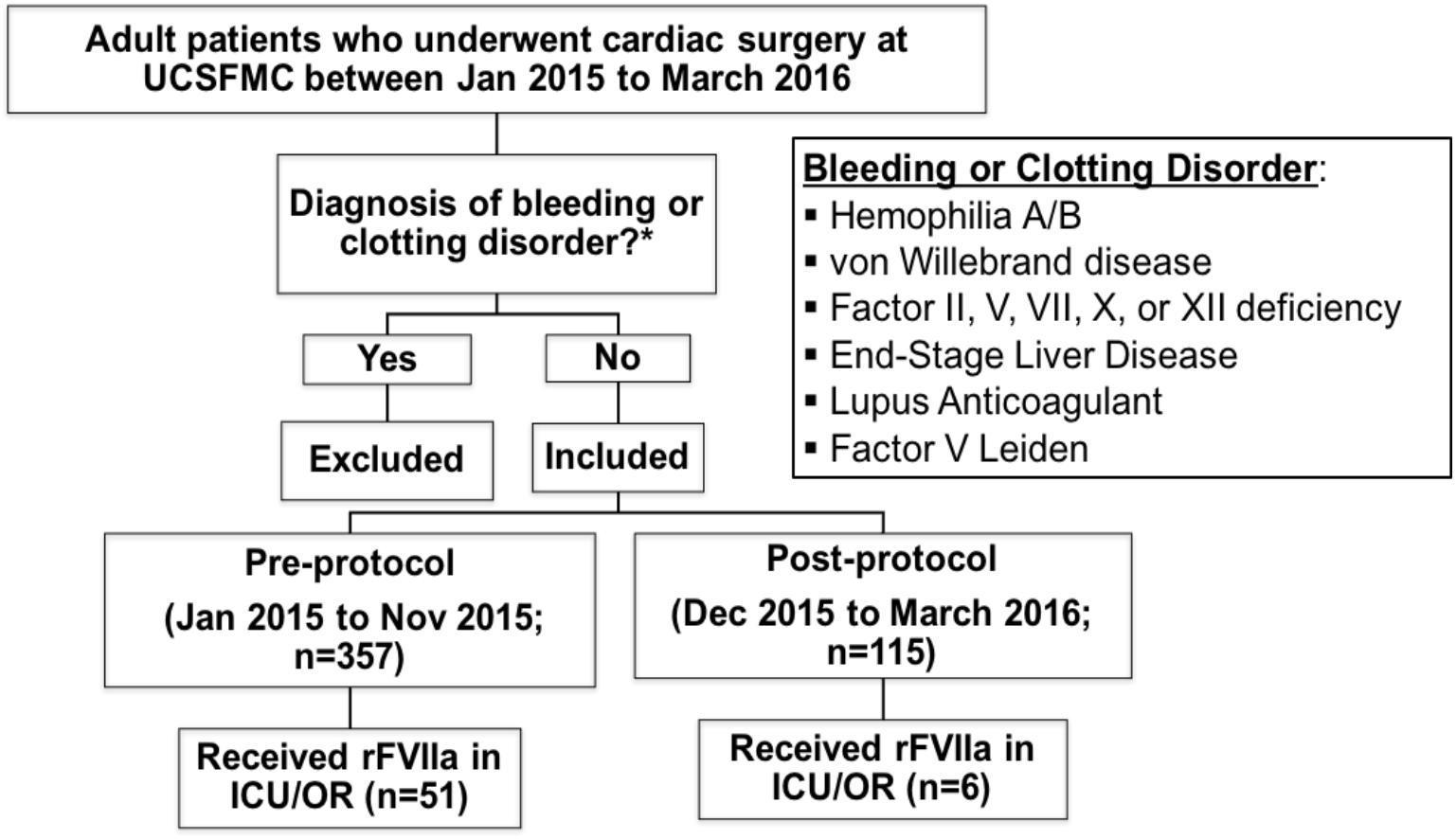
Study selection process pre- and post-protocol. ICU: intensive care unit, OR: operating room, UCSFMC: University of California, San Francisco, Medical Center.

**Table 1.** Baseline characteristics of pre- and post-protocol groups. Counts and percentages are reported for categorical data and means and standard deviations for continuous variables. AA: aminocaproic acid, BMI: body mass index, CABG: coronary artery bypass graft, ECMO: extracorporeal membrane oxygenation, SD: standard deviation, TXA: tranexamic acid, VAD: ventricular assist device.

|  | Pre-Protocol (n=51) | Post-Protocol (n=6) | p-value |
| --- | --- | --- | --- |
| **Average age±SD (years)** | 57.8±15.5 | 60.1±15.3 | 0.74 |
| **Male [n (%)]** | 40 (78) | 4 (66.7) | 0.89 |
| **Race [n (%)]** |  |  |  |
| Caucasian | 23 (45) | 4 (66.7) | 0.57 |
| Asian | 12 (23.5) | 1 (16.7) | >0.99 |
| African American | 6 (11.8) | 0 | 0.85 |
| Hispanic | 5 (9.8) | 1 (16.7) | >0.99 |
| Other/declined | 5 (9.8) | 0 | 0.97 |
| **Average BMI±SD (kg/m<sup>2</sup>)** | 25.4±5.5 | 26.2±4.4 | 0.73 |
| **Average weight±SD (kg)** | 75.1±18.9 | 83 ±22.0 | 0.43 |
| **Cardiac Procedure [n (%)]** |  |  |  |
| Valve replacement/repair | 25 (49.0) | 4 (66.7) | 0.70 |
| CABG | 12 (23.5) | 1 (16.7) | >0.99 |
| VAD | 5 (9.8) | 1 (16.7) | >0.99 |
| Heart transplant, ECMO,<br>or aortic dissection | 12 (23.5) | 1 (16.7) | >0.99 |
| **TXA or AA [n(%)]** | 44 (86.3) | 5 (83.3) | >0.99 |

### rFVIIa Use and Dosing

Use of rFVIIa significantly decreased post-implementation of our dosing standardization protocol targeting 30 mcg/kg (14.3% vs. 5.2%, p=0.015). Cardiac surgery patient cases were categorized based on the first rFVIIa dose they received (**Figure 2)**. Post-protocol, the percentage of cardiac surgery patients receiving ≤ 60 mcg/kg as their first dose was higher (83.3% vs. 45.1%, p=0.18) compared to the pre-protocol group. No post-protocol cases received a first dose in the 61-90 mcg/kg range compared to 39.2% of cases in the pre-protocol group (p=0.15). The percentage of cases that received a first dose greater than 90 mcg/kg during the pre- and post-protocol periods were not different (15.7% vs. 16.7%, p=0.60). Despite not reaching statistical significance due to the small sample size post-protocol, there was a clinically meaningful reduction in the average total dose per patient during the post-protocol period (81.4 vs. 56.6 mcg/kg, p = 0.059, **Table 2**). In addition, the location of the first dose administered shifted from the OR to the post-operative cardiac ICU setting; post-protocol, there was a decrease in the number of cases receiving rFVIIa in the OR (58.8% vs. 16.7%) and a rise in the number of doses administered in the ICU (41.1 vs. 83.3%), although this was not statistically significant.

**Figure 2.**
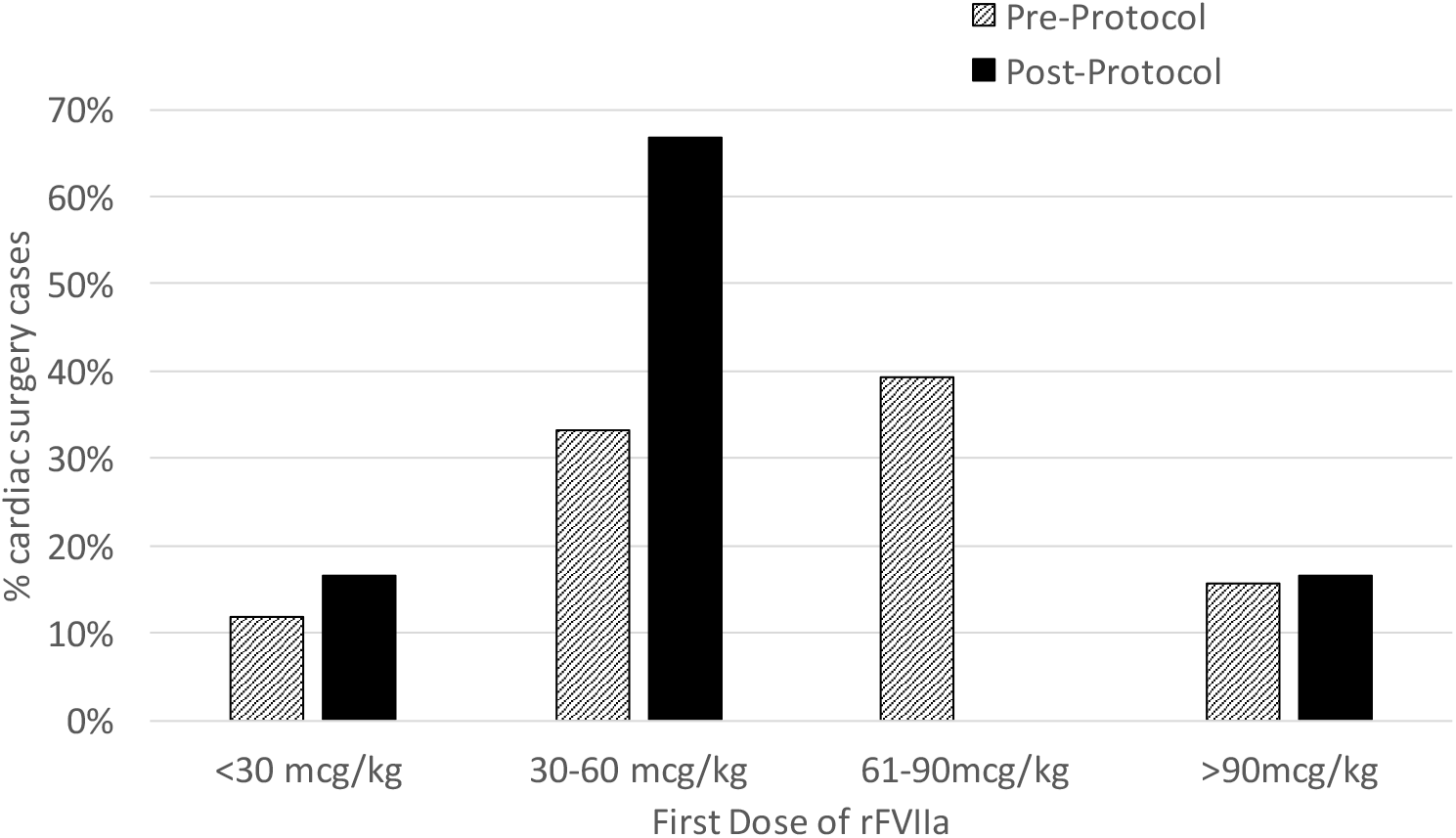
Distribution of first rFVIIa dose administered in cardiac surgery patients pre-vs. post-protocol. rFVIIA: recombinant factor VIIa.

**Table 2.** rFVIIa dosing characteristics and morbidity/mortality outcomes in cardiac surgery patients pre-vs. post-protocol. Counts and percentages are reported for categorical data and means and standard deviations for continuous variables. ED: emergency department, ICU: intensive care unit, OR: operating room, rFVIIA: recombinant factor VIIa.

|  | Pre-Protocol (n=51) | Post-Protocol (n=6) | p-value |
| --- | --- | --- | --- |
| **rFVIIa Dose (mcg/kg)** |  |  |  |
| Average Total Dose/patient | 81.4 | 56.6 | 0.059 |
| Average Dose/administration | 58.5 | 48.5 | 0.18 |
| Dose #1 | 62.9 | 54.7 | 0.27 |
| Dose #2 | 47.2 | 11.6 | 0.48 |
| Dose #3 | 46.4 | 0 | -- |
| **rFVIIa dosing range** | 19.1 – 135 mcg/kg | 11.6 - 130.4 mcg/kg | -- |
| **Patients who received >1 rFVIIa dose (%)** | 19 (37%) | 1 (16.7%) | 0.58 |
| **Location of 1<sup>st</sup> rFVIIa dose (n(%))** |  |  |  |
| OR | 30 (58.8%) | 1 (16.7%) | 0.13 |
| ICU | 21 (41.1%) | 5 (83.3%) | 0.13 |
| **Morbidity/Mortality Outcomes** |  |  |  |
| In-hospital mortality (n (%)) | 6 (11.8%) | 1 (16.7%) | >0.99 |
| Hospital length of stay (days) | 17.9 | 16.8 | 0.88 |
| 30-day need for re-exploration (n (%)) | 5 (9.8%) | 2 (33%) | 0.31 |
| 30-day readmission or ED visit (n (%)) | 7 (13.7%) | 1 (16.7%) | >0.99 |
| 30-day mortality (n(%)) | 0 | 0 | -- |

### Patient Outcomes

There were no significant differences in in-hospital mortality, hospital length of stay, 30-day need for re-exploration, 30-day readmission or ED visit, and 30-day mortality between pre- and post-protocol groups (**Table 2**).

### 4-Factor PCC Use and Drug Cost Analysis

While rFVIIa use declined, use of 4-factor PCC increased between the pre and post-protocol periods, i.e., 9/357 (2.5%) to 9/115 (8%), p=0.02 (**Figure 3A**). An average monthly total rFVIIa dose of 28 mg/patient was adminstered pre-protocol, and 6.8 mg/patient post-protocol. Using the average wholesale price (AWP) of $1.65/mcg for rFVIIa, we found the average monthly cost of rFVIIa significantly decreased from $45,900 to $11,138 (**Figure 3B**). After accounting for 4-factor PCC use, the total use of a factor VII-containing product (i.e., rFVIIa or 4-factor PCC) did not change (16.8% vs. 13%, p=0.416). An average monthly 4-factor PCC dose of 2,172 units/patient was used pre-protocol, and 4,284 units/patient post-protocol. The overall average cost per administration of either rFVIIa or 4-factor PCC (AWP

**Figure 3:**
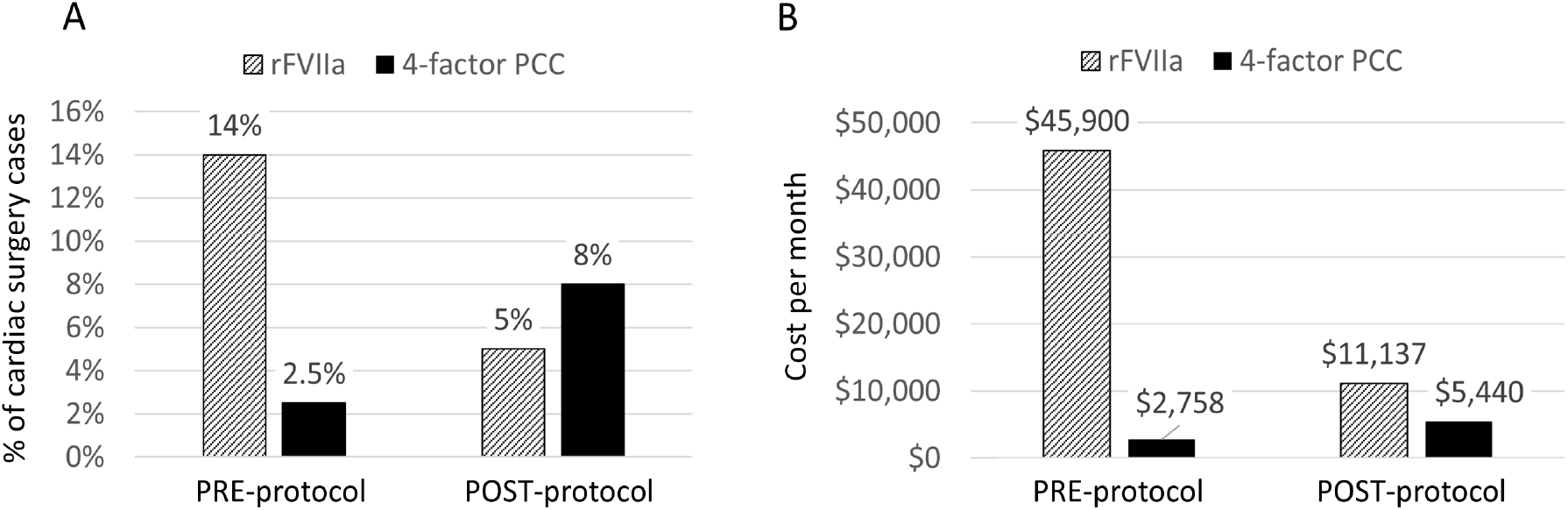
Cost analysis of rFVIIa and 4-factor PCC use between pre- and post-protocol periods. **(A)** Comparison of rFVIIa vs. 4-factor PCC use between pre- and post-protocol periods. **(B)** Comparison of rFVIIa and 4-factor PCC cost per month between pre- and post-protocol periods. PCC: prothrombin complex concentrate, rFVIIA: recombinant factor VIIa.

$1.27/unit) pre-protocol was significantly higher than the mean cost post-protocol ($8,778 vs. $4,421, p=0.00091). Despite the increased average cost per month of 4-factor PCC in the post-protocol period (from $2,758 to $5,440), the reduced cost of rVIIa resulted in an overall cost savings per month of approximately $32,000, or an estimated $385,000 annually.

## Discussion

Implementation of a low-dose rFVIIa pharmacist stewardship program in cardiac surgery at our institution effectively reduced the overall use and dose of rFVIIa, which equated to drug cost savings. Few studies have been published on outcomes of institutional rFVIIa stewardship protocols, and our study is unique in that no others have investigated the use of 4-factor PCC in response to a restriction in rFVIIa dosing in cardiac surgery.

Our results are comparable to other studies that implemented rFVIIa stewardship protocols addressing off-label use; available studies are observational, retrospective and have small sample sizes. In 2008, Owen *et al*. studied the effect of a guideline directing appropriate use and dose of rFVIIa for multiple off-label uses including neurosurgery, CT surgery, and liver transplant.^25^ There was a signficant reduction in mean total dose per patient from 81.1 mcg/kg to 45.3 mcg/kg and semiannual cost savings by $110, 014.^25^ Similar to our work, there were no changes in adverse patient outcomes.^25^ Bain *et al* (2014) found that a pharmacist-driven rFVIIa protocol targeting 30 mcg/kg/dose in critical bleeding for a variety of off-label indications signficantly reduced the dose (47.5 vs. 62.2 mcg/kg) with no change in mortality.^26^ In 2015, Trueg *et al* found a 32% reduction in rFVIIa use and an annual cost savings of $375,539, with no difference in mortality; this study was not restricted to cardiac surgery, and the majority of patients had an underlying history of liver disease.^27^ Marsh *et al*. in 2020 found a reduction of $414,500 in annual costs of rFVIIa after transitioning rFVIIa from the blood bank to pharmacy and implementing a dosing guide (i.e. 20 mcg/kg in cardiac surgery).^28^ The study was not restricted to cardiac surgery and included patients with approved indications (e.g., factor VII deficiency).^28^ Lastly, Waheed *et al*. in 2020 investigated pre- and post-protocol changes with the involvement of a hematology consult to guide the appropriate use of off-label 4-factor PPC and rFVIIa.^29^ Although use before and after stewardship was similar, there was a total semiannual cost savings of $101,736.^29^

There were several limitations to this study. The post-protocol period of four months resulted in a small sample size. In addition, although the protocol was approved in December 2015, several weeks were required to adequately train the OR and ICU pharmacists responsible for verifying the majority of rFVIIa orders. Additionally, while most of the surgeons were eager to comply with the weight-based dosing recommended in the protocol, there was inter-prescriber variability in rFVIIa doses ordered. These reasons may have contributed to our inability to decrease the average post-protocol dose to the 30 mcg/kg recommendation. This dosing protocol was part of a larger algorithm to address microvascular bleeding during cardiac surgery, which instructs prescribers to use other blood products before ordering rFVIIa. Our initial study design intended to describe the use of rFVIIa with respect to the use of other blood products; however, since blood product use is recorded in a separate medical record documentation system not accessible to pharmacy, we were unable to collect these data nor understand where rFVIIa falls with respect to overall use of blood products. Lastly, our 30-day outcomes may have been limited by a lack of integration of electronic medical records. Since UCSF is a tertiary care referral center, many patients do not receive follow-up care at UCSF, so readmission and mortality data were limited.

There was an unexpected outcome of our stewardship protocol—a concomitant, significant increase in 4-factor PCC use for cardiac surgery bleeding. The off-label use of PCC has expanded recently to a number of indications, including cardiac surgery populations.^30^ Given the small volume of concentrated clotting factors, it may be beneficial over large-volume blood products in the cardiac population and has been shown to reduce post-operative blood loss and RBC transfusion requirements compared to FFP in cardiac surgery.^31^ There is limited guidance in 4-factor PCC dosing for refractory bleeding in cardiac surgery and there are no large randomized controlled trials in this area. More recently, we have learned low-doses of 4-factor PCC may be sufficient; a 2021 study found low-dose rFVIIa (2 mg or 20 mcg/kg) and low-dose 4-factor PCC (1000 units or 15 units/kg) were equally safe and effective for refractory bleeding in cardiac surgery.^32^ In the absence of a 4-factor PCC dosing protocol, there is potential for administration of larger than necessary doses (e.g., 2,172 units/patient pre-protocol and 4,284 units/patient post-protocol in our study). Therefore, if restricting rFVIIa off-label use, it seems critical to also standardize use and dosing of both PCC and rFVIIa to avoid shifts in prescribing or overutilization of PCC.

## Conclusions

The use of rFVIIa for bleeding during cardiac surgery decreased after implementation of a rFVIIa protocol targeting 30 mcg/kg/dose without compromising morbidity or mortality outcomes. Additionally, the length of stay, need for re-exploration, and readmission rates were not compromised. Although 4-four-factor PCC significantly increased post-protocol, use of rFVIIa or 4-factor PCC did not change.

Decreased rFVIIa use resulted in significant cost savings, which were maintained after accounting for increased 4-factor PCC use. As 4-factor PCC use increased significantly during the study period, institutions wanting to implement a rFVIIa protocol should take measures to concurrently address off-label use of 4-factor PCC.

## Data Availability

All data produced in the present study are available upon reasonable request to the authors

## Acknowledgements

We thank Dr. James Ramsay for his input.

## Conflict of Interest Statement

None of the authors have any disclosures, conflicts of interest, or sources of funding for this work.

## Notes

### Competing Interest Statement

The authors have declared no competing interest.

### Funding Statement

This study did not receive any funding

### Author Declarations

Institutional review board (IRB) approval was obtained for this study through the University of California, San Francisco, Committee on Human Research Protection Program.

